# Low to moderate genetic influences on the rapid smell test SCENTinel™

**DOI:** 10.1101/2023.05.14.23289965

**Authors:** Stephanie R. Hunter, Cailu Lin, Mackenzie E. Hannum, Katherine Bell, Amy Huang, Paule V. Joseph, Valentina Parma, Pamela H. Dalton, Danielle R. Reed

## Abstract

SCENTinel^™^ - a rapid, inexpensive smell test that measures odor detection, intensity, identification, and pleasantness - was developed for population-wide screening of smell function. SCENTinel^™^ was previously found to screen for multiple types of smell disorders. However, the effect of genetic variability on SCENTinel^™^ test performance is unknown, which could affect the test’s validity. This study assessed performance of SCENTinel^™^ in a large group of individuals with a normal sense of smell to determine the test-retest reliability and the heritability of SCENTinel^™^ test performance. One thousand participants (36 [IQR 26-52] years old, 72% female, 80% white) completed a SCENTinel^™^ test at the 2021 and 2022 Twins Days Festivals in Twinsburg, OH, and 118 of those completed a SCENTinel^™^ test on each of the festival’s two days. Participants comprised 55% percent monozygotic twins, 13% dizygotic twins, 0.4% triplets, and 36% singletons. We found that 97% of participants passed the SCENTinel^™^ test. Test-retest reliability ranged from 0.57 to 0.71 for SCENTinel^™^ subtests. Broad-sense heritability, based on 246 monozygotic and 62 dizygotic twin dyads, was low for odor intensity (r=0.03) and moderate for odor pleasantness (r=0.4). Together, this study suggests that SCENTinel^™^ is a reliable smell test with only moderate heritability effects, which further supports its utility for population-wide screening for smell function.

## Introduction

The COVID-19 pandemic has highlighted the need for a rapid, inexpensive, easily administered smell test to help screen for smell loss throughout the population^1–7^. Besides COVID-19 symptom screening, such a smell test can be useful in many other areas where smell loss is a sign or symptom, including early identification of neurodegenerative diseases^8^, 5-year mortality rate^9–15^, and severity of traumatic brain injury^16^. Furthermore, regularly testing smell function can help identify individuals who may unknowingly suffer from smell loss^5^, such as those who were born without a sense of smell or who experience gradual smell loss due to aging or viral illnesses, and implement lifestyle changes to prevent potential hazards or begin therapies.

However, for a smell test to be useful in these situations, it must be accurate and reliable. Even within a group of people with a normal sense of smell, several individual differences may affect smell function, which a smell test must consider. For example, women typically outperform men in terms of odor identification, odor discrimination, and odor threshold abilities^17^. Smell function also declines during normal aging^18^; most people over the age of 80 have smell impairment^19^, with some odors more impaired with age than others^20^. Genetic differences in olfactory receptors can impact the perceived intensity and pleasantness of an odor^21^, in addition to whether someone can smell a particular odor or not (i.e., specific anosmia). The most notable example of this is the odorant androstenone, which smells unpleasant to some but others cannot smell it at all^22^. A smell test that includes an odor for which part of the population has a specific anosmia, due to their genetic makeup, may impair the test’s utility in population-wide screening for smell function. Measuring the effects of heritability in smell test responses can determine if it contributes to person-to-person variation, which would reduce the accuracy of such a smell test.

In 2020 we created SCENTinel^™^, a rapid, inexpensive, easily administered smell test to help screen for smell disorders throughout the population. SCENTinel^™^ uses three Liftn’Smell labels (Scentisphere LLC, Caramel, NY), two of which are blank and one has an odor, to measure odor detection, odor intensity, odor identification, and odor pleasantness, providing a cumulative overall score to determine one’s smell function. The first version of the test (SCENTinel^™^ 1.0), which included one odor option (flower), was found to distinguish people with anosmia from those with normosmia and was validated against the NIH Toolbox Odor Identification Test^7^. The second version of the test (SCENTinel^™^ 1.1) included one of four possible odors (flower, coffee, bubblegum, caramel popcorn) and was found to screen for individuals with anosmia, hyposmia, parosmia, and normosmia^23^. The version used in this study (SCENTinel^™^ 2.1) expands on the previous studies and includes one of nine possible odors (flower, coffee, bubblegum, orange, strawberry, banana, coconut, woody, and lemon). This study was the first time SCENTinel^™^ 2.1 was tested, and it was still unknown whether these odor versions performed similarly. Furthermore, whether heritability affected SCENTinel^™^ test performance and whether SCENTinel^™^ is reliable over time were also unknown.

Understanding how each of these factors could affect SCENTinel^™^ test performance is necessary to assess SCENTinel’s^™^ accuracy and reliability as a population-wide smell test. The purpose of this study was to assess performance of SCENTinel^™^ 2.1 in a large group of individuals with normosmia to determine test-retest reliability and heritability of SCENTinel^™^ 2.1 subtests and overall score.

## Materials and methods

### Participants

Participants 18 years or more of age were recruited at the Twins Days Festivals in Twinsburg, Ohio, which took place on August 7 and 8, 2021, and August 6 and 7, 2022. The festival, which celebrates the uniqueness of twins and others of multiple births, is attended by twins and their families from around the world. Overall, 1,078 participants enrolled in this study. We removed participants with a known, preexisting smell or taste disorder (n=13), who did not complete the entire SCENTinel^™^ test (n=17), or who were found to be given an incorrectly labeled SCENTinel^™^ card (n=48). Thus, 1,000 participants (72% female, 80% white, median age 36 [IQR 26-52] years old; Table 1) were included in the final analyses, comprising 128 individuals who were part of a dizygotic twin pair, 498 individuals who were part of a monozygotic twin pair, and four individuals who were part of a triplet; the remaining 362 participants were singletons (i.e., not a part of a twin or triplet). Of these, 118 participants (69% female, 89% white, median age = 38 [IQR 25-54] years; Supplementary Table S1) took the test on both days of the festival and were included in the test-retest analysis (Supplementary Table S1).

**Table 1.** Participant Demographics.

|  | **N** | **%** |
| --- | --- | --- |
| **Total subjects** | **1000** | **100** |
| **Age (median [IQR] years)** | **36 [26-52]** |  |
| Young (18-41) | 592 | 60 |
| Middle (42-63) | 287 | 29 |
| Old (64-88) | 101 | 10 |
| Declined to provide | 20 | 2 |
| **Sex** |  |  |
| Female | 715 | 72 |
| Male | 279 | 28 |
| Other | 2 | 0.2 |
| Prefer not to say | 4 | 0.4 |
| **Race/Ethnicity** |  |  |
| White | 804 | 80 |
| Black or African American | 107 | 11 |
| Asian (including South Asian and Asian Indian) | 23 | 2.3 |
| Hispanic or Latino | 30 | 3 |
| American Indian or Alaska Native | 7 | 0.7 |
| Native Hawaiian or Other Pacific Islander | 3 | 0.3 |
| Other | 19 | 1.9 |
| Prefer not to say | 7 | 0.7 |
| **Twin status** |  |  |
| Monozygote | 498 | 50 |
| Dizygote | 128 | 13 |
| Triplet | 4 | 0.4 |
| Singleton | 362 | 36 |
| Unknown | 8 | 0.8 |

### SCENTinel^™^ 2.1

The SCENTinel^™^ test has two components: the test card, where participants smell the odor, and the online survey, where participants record their answers. The SCENTinel^™^ test card contains three Liftn’Smell labels (Scentisphere LLC, Caramel, NY), labeled A, B, and C. Only one of these labels contains a target odor; the other two are blank. SCENTinel^™^ 2.1 uses one of nine target odorants (flower, bubblegum, orange, coffee, banana, strawberry, coconut, woody, or lemon), which can be located under any one of the three labels on the SCENTinel^™^ test. The odorants, from Givaudan and Robertet (for catalog numbers, see Supplementary Table S2), were designed to be iso-intense, at an intensity of 80 on a scale of 0 to 100, confirmed via pilot testing.

SCENTinel^™^ is completed using an online REDCap survey^24^, which can be accessed through a QR code or through a URL, both on the SCENTinel^™^ test card. SCENTinel^™^ tests four olfactory measures: odor detection accuracy (correct/incorrect); odor intensity (from 1 to 100, treated as a dichotomous [above/below a cutoff of 20] or a continuous variable, depending on the analysis); odor identification among four given options (i.e. one correct response and three distractors; correct/incorrect) or, if the first response is incorrect, among the three remaining options (i.e. one correct response and two distractors; correct/incorrect); and odor pleasantness (from 1 to 100, continuous value). Odor detection, odor intensity, and odor identification were used to determine an overall score of pass or fail (see Supplementary Table S3).

### Procedure

This study was approved by the University of Pennsylvania Institutional Review Board (protocol no. 844425) and complied with the Declaration of Helsinki. In this festival setting, all testing was done under a tent outside at ambient temperature. Interested participants were provided a unique ID number and given a SCENTinel^™^ test card, which they completed while seated at tables under the tent, using their smartphone or, if they did not have one, using a provided iPad. Researchers were available if participants had questions or technical issues. Twin dyads or triplet triads, unbeknownst to them, were given a SCENTinel^™^ test with the same odor, but the odor could be under different labels. Twins who were tested at the same time were asked not to sit next to each other, to avoid sharing answers during the test. Participants accessed the online REDCap survey^24^ through a QR code or a URL located on the SCENTinel^™^ test. They first provided consent using the approved online consent form and then provided demographic information (age, sex, race, and their Twins participant ID number, if applicable) and indicated whether they had issues with their taste and smell. Participants were then instructed to lift and smell each label on the SCENTinel^™^ card, one at a time, from left to right, and 1) select which label smelled the strongest (odor detection), 2) rate how intense the odor was on a 100-point visual analog scale anchored with “No Smell” at 0 and “Very Strong Smell” at 100 (odor intensity), and 3) identify the odor out of four possible options (odor identification). If they identified the odor incorrectly the first time, they were given a second chance (three-alternative forced-choice paradigm). Binary responses to these subtests (correct/incorrect, with an odor intensity cutoff of 20 out of 100) were used to calculate participants’ overall score (pass/fail), as previously specified^7^. Participants then rated the pleasantness of the odor on the SCENTinel^™^ test card using a 100-point visual analog scale anchored with “Very Unpleasant” at 0 and “Very Pleasant” at 100.

All participants were invited to participate in this study both days of the festival, to assess test-retest reliability, following the same procedure described above. Unbeknownst to them, participants who returned on the second day were provided with a SCENTinel^™^ test that had the same odor as their previous test, but possibly under a different label.

### Statistical Analysis

All data analysis was performed in R using R Studio software (version 2022.12.0+353). Age was categorized such that the age span was equal for young, middle, and old age categories. For all analyses except test-retest, we included only participants’ first test results.

Multiple linear regression was used to determine the effects of age, sex, race, and target odor on continuous variables (i.e., odor intensity and odor pleasantness). When main effects were significant (p<0.05), pairwise comparisons were assessed with Tukey post-hoc tests. Binary logistic regression was used to determine the effects of age, sex, race, and target odor on binary categorical variables (i.e., odor detection, odor identification, overall score). The Wald test was used to determine the overall effect of coefficients. Pearson and tetrachoric correlation coefficients were used to determine test-retest reliability for continuous and binary categorical variables, respectively. P < 0.05 was considered significant.

Heritability was calculated using data from all twin pair participants (monozygotic twin pairs, 80.2%; dizygotic twin pairs, 19.8%), using structural equation methods^25^ that partition the variation of quantitative variables into variance attributable to specific effects: additive genetic (A), dominant genetic (D), shared environmental (C), and unique environmental (E). These variance components were decomposed in a polygenic model by specific type of analyses, or modes (ACE, ADE, AE, CE, and E), computed with *twinlm* function^26^. Heritability of binary categorical variables was calculated using a liability-threshold model by defining variance components similarly as quantitative variables and was computed with *bptwin* function^27^. The estimation of genetic heritability was performed using the *mets* package (version 1.3.2) of R statistical software.

## Results

### Effects of age, sex, and race on SCENTinel^™^ performance

Most participants passed the odor detection (96%), odor intensity (99.7%), odor identification (first attempt, 76%; second attempt, 59%), and SCENTinel^™^ overall score (97%; Table 2). The average odor intensity rating was 79 ± 14. Participants overall rated the odors as pleasant (mean ± SD rating, 75 ± 22 out of 100). Odor intensity ratings showed significant effects of age [F (2,954) = 9.8; p < 0.001] and sex [F (1,954 = 8.4; p = 0.004; Figure 1]. Females (79 ± 14 out of 100) and young (18 - 41 years old) participants (80 ± 13) rated the odor as more intense than did males (77 ± 15; p = 0.002) and middle-aged (42 - 63 years; 77 ± 15; p = 0.005) and older (64 - 88 years; 75 ± 17; p < 0.001) participants, respectively. White participants had higher odds of correctly detecting the odor on the SCENTinel^™^ test (96% accurate) compared to nonwhite participants (93% accurate; OR 2.08, 95% CI [0.02, 1.4]). Furthermore, older participants (64 - 88 years) had a lower odds of accurately identifying the target odor on the first attempt (63% accurate; OR 0.42, 95% CI [0.25, 0.68], p < 0.001) than did young participants (18 - 41 years; 78% accurate; Table 3).

**Table 2.**
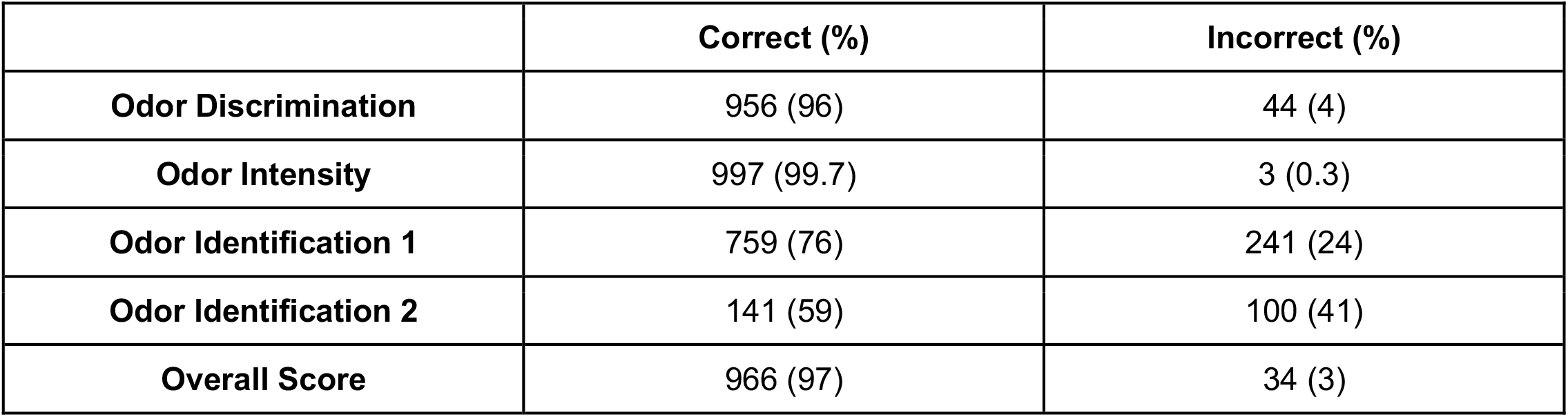
Performance on SCENTinel^™^ 2.1 subtests and overall performance.

**Table 3.** Differences in age, sex, race, and target odor for odor identification accuracy. Odds ratios (ORs), 95% confidence intervals (95% CIs). References: age = young; sex = female; race = nonwhite; target odor = flower.

| Variable | OR | 95% CI | p-Value |
| --- | --- | --- | --- |
| Age - Middle | 0.88 | 0.61, 1.26 | 0.47 |
| Age - Old | 0.42 | 0.25, 0.68 | <0.001 |
| Sex - Male | 0.91 | 0.65, 1.29 | 0.60 |
| Race - White | 1.47 | 0.99, 2.19 | 0.054 |
| Odor - Bubblegum | 0.38 | 0.1, 1.2 | 0.12 |
| Odor - Woody | 0.49 | 0.13, 1.59 | 0.26 |
| Odor - Coconut | 0.15 | 0.04, 0.41 | <0.001 |
| Odor - Coffee | 0.14 | 0.04, 0.38 | <0.001 |
| Odor - Lemon | 0.14 | 0.04, 0.42 | <0.001 |
| Odor - Orange | 0.1 | 0.03, 0.28 | <0.001 |
| Odor - Strawberry | 0.05 | 0.02, 0.14 | <0.001 |
| Odor - Banana | 0.04 | 0.01, 0.11 | <0.001 |

**Figure 1.**
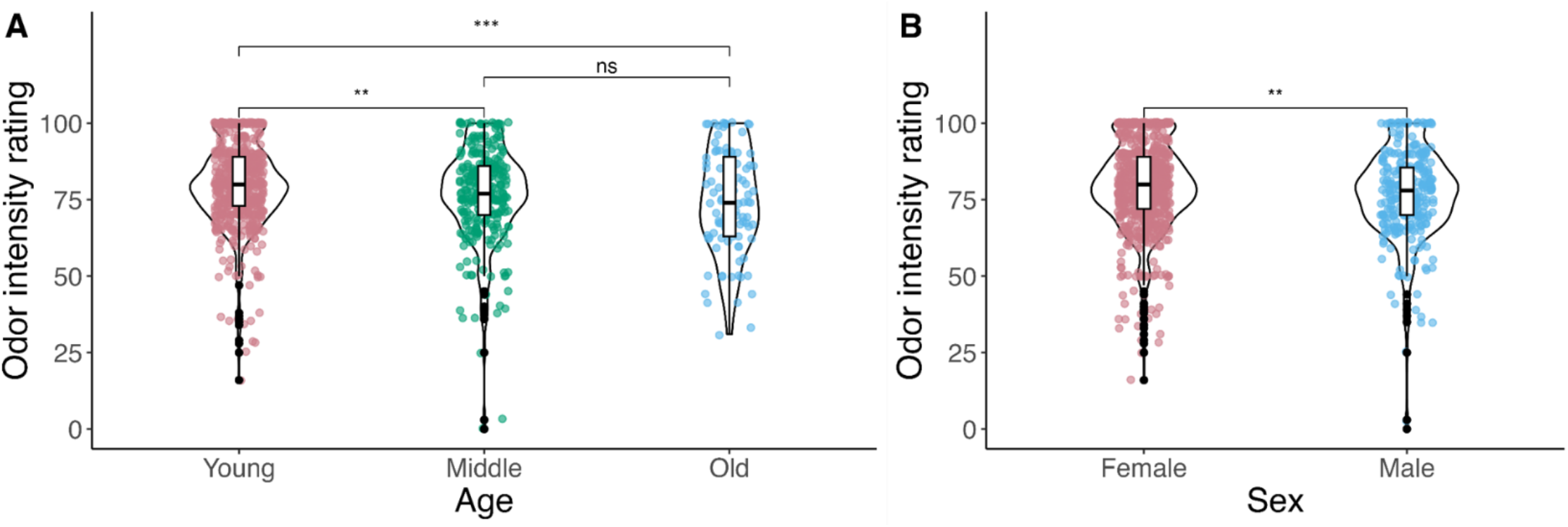
Significant effects of age (A) and sex (B) on SCENTinel^™^ odor intensity ratings. **p<0.01; ***p<0.001; ns = not significant.

### Differences in SCENTinel^™^ performance between the nine possible odors

The nine odors used across SCENTinel^™^ tests did not differ in pass rate for odor detection (χ^2^ = 4.9, df = 8, p = 0.77) or in odor intensity ratings (as continuous variable; F(8,954) = 1.34; p = 0.2). However, they did differ in odor identification accuracy (χ^2^ = 96, df = 8, p < 0.001), as follows (Figure 2): flower (95%), bubblegum (91%), woody (90%), coconut (77%), coffee (77%), lemon (76%), orange (73%), strawberry (57%), and banana (53%). For those who misidentified banana, the most selected distractor was flower (34%) [other distractors included lemon (9.4%) and natural gas (3.9%)]. As shown in Table 3, compared to the target odor flower, the target odors coconut (OR 0.15, 95% CI [0.04, 0.41]), coffee (OR 0.14, 95% CI [0.04, 0.38]), lemon (OR 0.14, 95% CI [0.04, 0.42]), orange (OR 0.1, 95% CI [0.03, 0.28]), strawberry (OR 0.15, 95% CI [0.04, 0.41]), and banana (OR 0.04, 95% CI [0.01, 0.11]) all had significantly (p < 0.001) lower odds of being identified correctly. The nine odors also differed in pleasantness ratings (F(8, 952) = 6.4; p < 0.001). Figure 3 shows that the woody odor (65 ± 26) was rated as significantly less pleasant than lemon (83 ± 18; p < 0.001), orange (81 ± 18; p < 0.001), strawberry (77 ± 18; p = 0.002), bubblegum (76 ± 23; p = 0.02), banana (76 ± 21; p = 0.003), and coconut (75 ± 21; p = 0.02). The coffee odor (69 ± 25) was rated significantly less pleasant than lemon (p < 0.001) or orange (p < 0.001). Lastly, the nine odors used across SCENTinel^™^ tests did not differ in passing rate for overall score (χ^2^ = 4.6, df = 8, p = 0.8): 100% of those with the woody odor passed, 98% of those with the flower or coconut odor passed, 97% of those with the banana or lemon odor passed, 96% of those with the coffee odor passed, 95% of those with the orange passed, and 94% of those with the bubblegum or strawberry odors passed the overall score.

**Figure 2.**
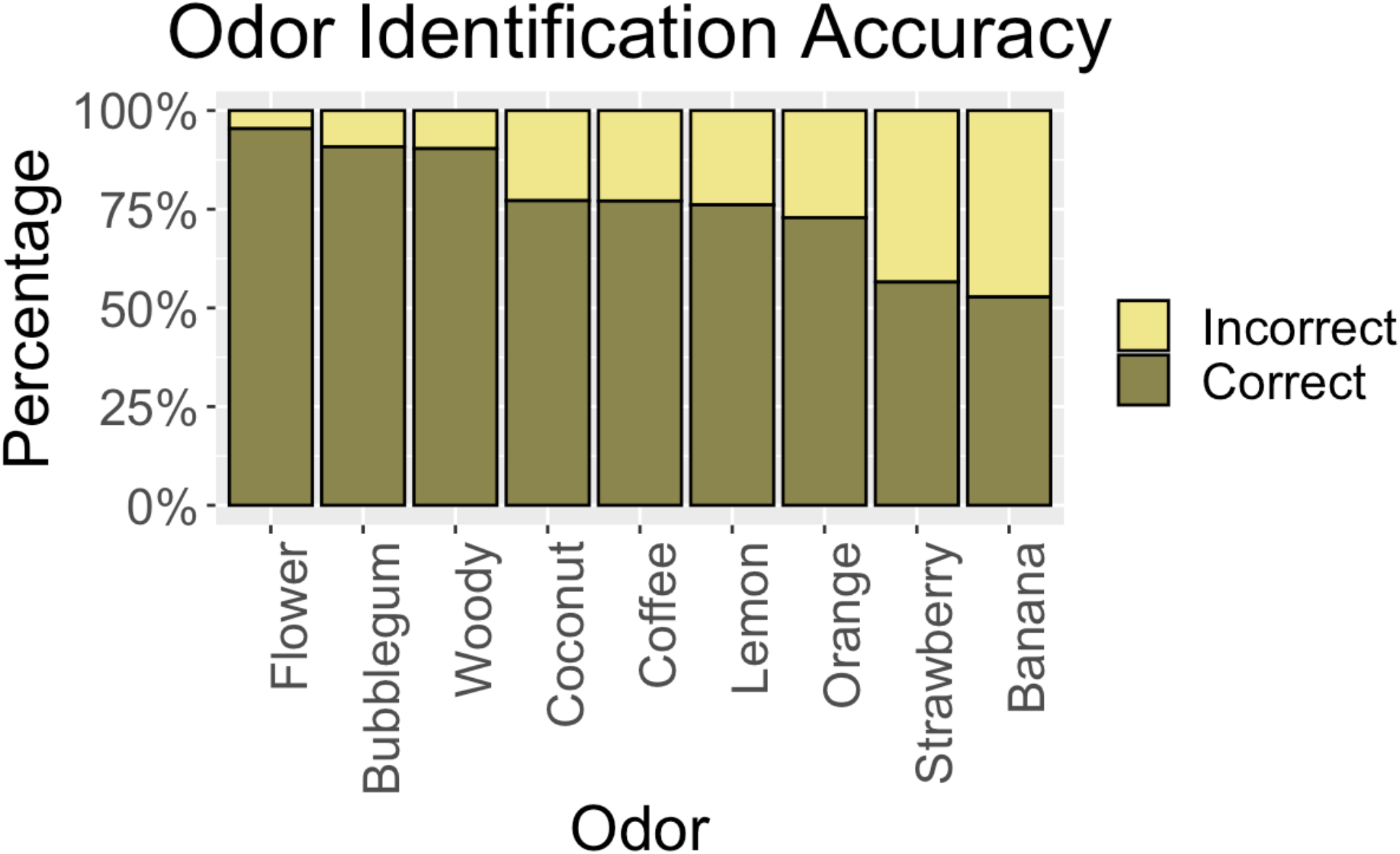
Odor Identification accuracy (first attempt) across all nine target odors.

**Figure 3.**
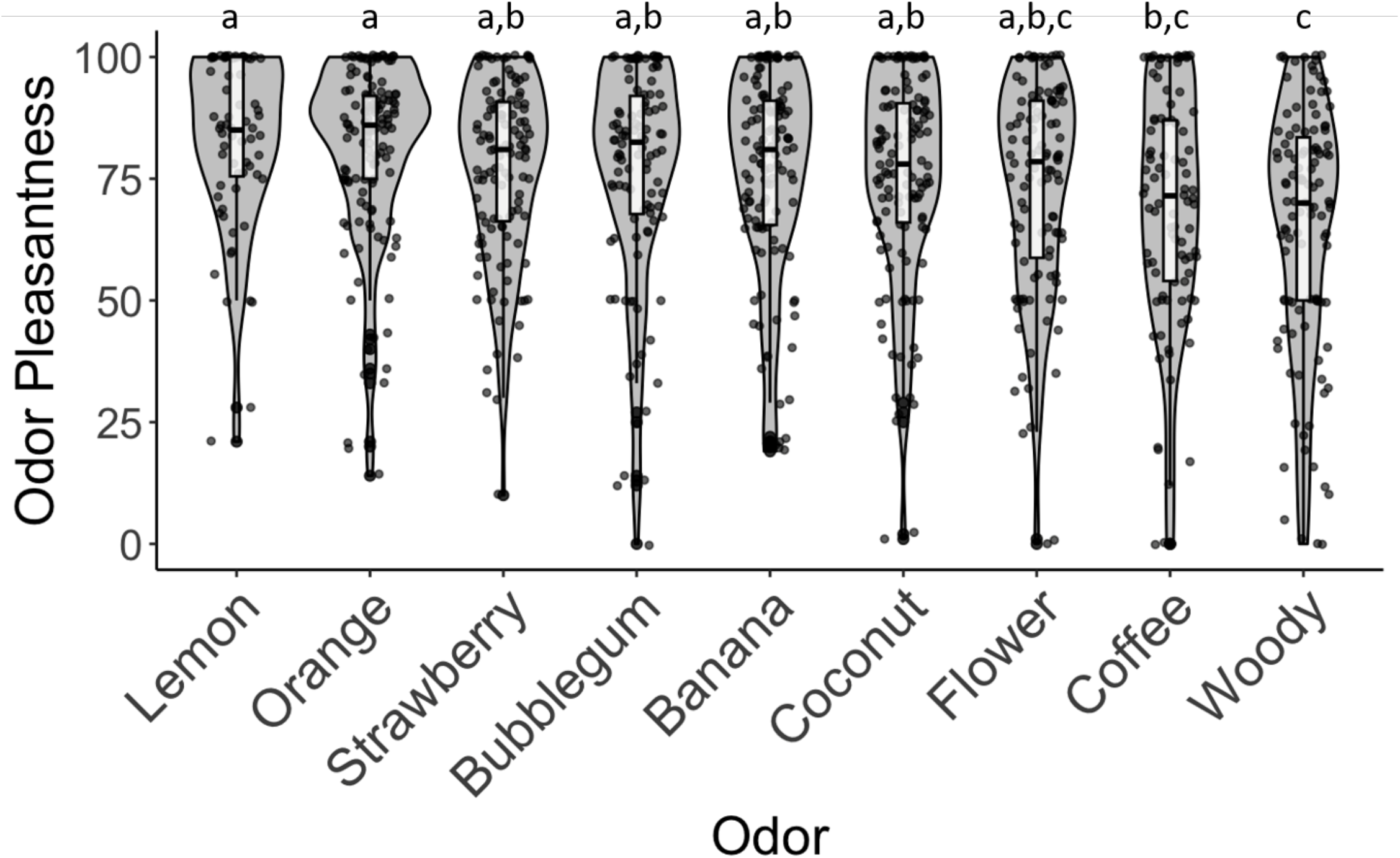
Odor pleasantness ratings for each of the nine target odors used across SCENTinel^™^ tests. Dots represent individual responses.

### Test-Retest

The test-retest correlation coefficients for odor intensity (continuous variable; r = 0.35, p < 0.001), odor identification (r = 0.71; p < 0.001), and odor pleasantness (r = 0.61, p < 0.001) were all moderate to strong (Figure 4; Table 4). The low test-retest reliability for Odor Identification 2 is likely due to the small sample size that needed a second attempt. Overall, among participants who took the test on both days, 97% answered odor detection correctly, 99% answered odor intensity correctly (i.e., >20/100), and 97% had passing SCENTinel^™^ overall scores on both days. Among these, 79% correctly and 5% incorrectly identified the target odor both days (total concordance is 84%). Of participants who incorrectly identified the target odor on the first attempt both days, 17% answered correctly on the second attempt both days, and of participants who incorrectly identified the target odor on the first attempt both days, 33% answered incorrectly on the second attempt both days (total concordance is 50%).

**Table 4.**
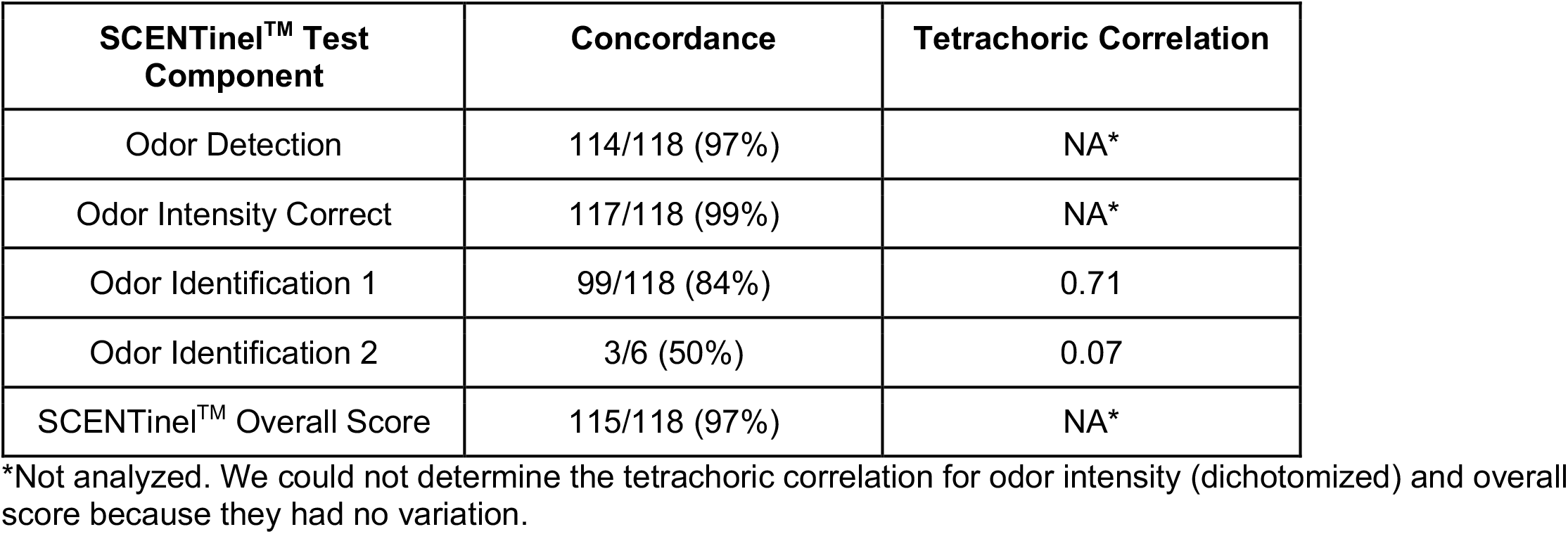
Test-retest reliability for odor discrimination, odor intensity, odor identification, and SCENTinel^™^ overall score.

| SCENTinel™ Test Component | Concordance | Tetrachoric Correlation |
| --- | --- | --- |
| Odor Detection | 114/118 (97%) | NA* |
| Odor Intensity Correct | 117/118 (99%) | NA* |
| Odor Identification 1 | 99/118 (84%) | 0.71 |
| Odor Identification 2 | 3/6 (50%) | 0.07 |
| SCENTinel™ Overall Score | 115/118 (97%) | NA* |

\*Not analyzed. We could not determine the tetrachoric correlation for odor intensity (dichotomized) and overall score because they had no variation.

**Figure 4.**
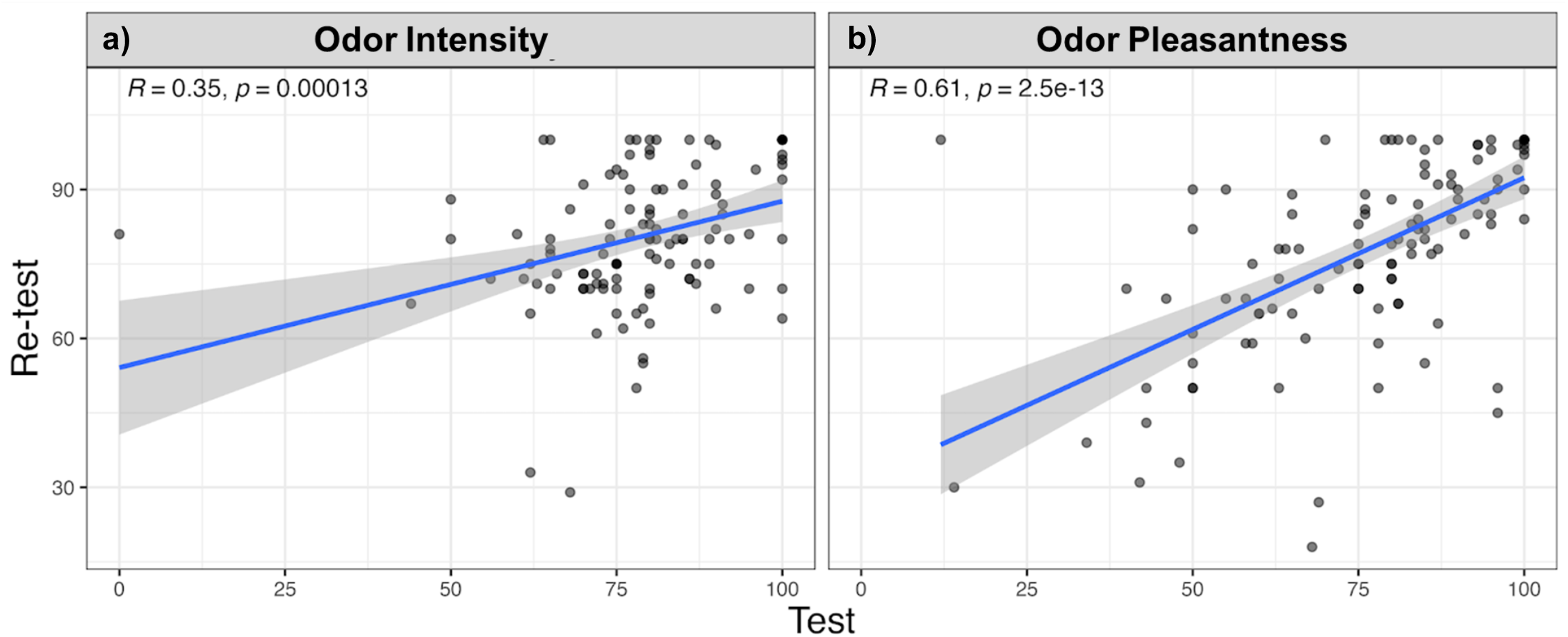
Test-retest reliability for a) odor intensity and b) odor pleasantness.

### Heritability

A small number of participants failed odor detection, odor identification, and the SCENTinel^™^ overall score, creating unbalanced data, so we assessed heritability only for continuous variables (for full results, see Supplementary Table S4). Overall, monozygotic twins had stronger correlations than did dizygotic twins (Table 5). Broad-sense heritability is estimated at 0.03 [95% CI (−0.09 to 0.15)] for odor intensity and 0.40 [95% CI (0.3 - 0.5)] for odor pleasantness. Thus, the heritability for odor intensity is low, while the heritability for odor pleasantness is moderate. However, the odors were designed to be rated at 80 on a 0-100 scale and were considered intense by most of the participants. Thus, there may be a ceiling effect that prevented us from detecting any differences in odor intensity ratings due to heritability.

**Table 5.** Heritability among monozygotic (MZ) and dizygotic (DZ) twins.

| SCENTinel™<br>Test<br>Component | N |  | Pearson's Correlation<br>(95% CI) |  | Broad-Sense<br>Heritability<br>(95% CI) |
| --- | --- | --- | --- | --- | --- |
|  | MZ | DZ | MZ | DZ |  |
| Odor Intensity | 246 | 62 | 0.03<br>(-0.09 to 0.14) | 0.02<br>(-0.04 to 0.07) | 0.03<br>(-0.09 to 0.15) |
| Odor<br>Pleasantness | 246 | 62 | 0.40<br>(0.3 - 0.5) | 0.20<br>(0.2 - 0.3) | 0.40<br>(0.3 - 0.5) |

## Discussion

The COVID-19 pandemic highlighted the need for population-wide screening of smell function to monitor disease and health status^6,7^. The SCENTinel^™^ rapid smell test, created to fill this gap^7^, is an inexpensive, quick, self-administered smell test that has previously been found to screen for normosmia, hyposmia, anosmia, and parosmia^23^. For SCENTinel^™^ to monitor smell function throughout the population, it must be accurate and reliable and not prone to failure due to genetic differences in smell function (e.g., specific anosmias). This study assessed performance of SCENTinel^™^ in a large group of normosmics to determine test-retest reliability and heritability of SCENTinel^™^ scores.

Overall, 97% of participants passed the SCENTinel^™^ rapid smell test, as expected for self-reported normosmics. However, among the subtests, different odors had different rates of correct identification. When developing the test, our goal was to select odors and distractors such that 80% of people with a normal sense of smell can correctly identify the correct odor. In this SCENTinel^™^ version, however, coconut, coffee, lemon, orange, strawberry, and banana were correctly identified less than 80% of the time. The odors used in SCENTinel^™^ were chosen because they are highly familiar to adults in the United States^28–30^, making it unlikely that the odors were unrecognizable. Thus, the odor distractors may have been too similar to the target odor, reducing identification accuracy^31^. After this study, the odor distractors on SCENTinel^™^ were changed so that they better contrasted with the correct odor. For example, in this version of SCENTinel^™^ banana was most often incorrectly identified as flower; after this study, flower was removed as a distractor and replaced with popcorn.

In addition to the majority of normosmics passing the SCENTinel^™^ test, SCENTinel^™^ also had moderate to high test-retest reliability for all components, indicating that it is not only accurate but also reliable. The test-retest reliability for the gold-standard smell tests UPSIT (40 items) and Sniffin’ Sticks extended version (16-item identification test plus discrimination and threshold tests) is 0.95 and 0.93^32^, respectively, but these tests require a lot of time to complete due to the extensive list of odors used. In general, when fewer odorants are used, reliability tends to decrease. For example, the test-retest reliability of the identification component of the 16-odor version of Sniffin’ Sticks is 0.73 and falls to 0.71 when 12 odors are used per test^33^, and when the UPSIT is reduced to 12 odors (i.e. in the CC-SIT), its test-retest reliability is also 0.71^34^. SCENTinel^™^, despite using only a single odor per test, also has a test-retest correlation of 0.71 for the identification subtest, making this much shorter test comparable in reliability.

The differences in SCENTinel^™^ performance across age and sex groups match findings from previous studies. It is well documented that women have higher olfactory sensitivity than do men^17,35^, and previous studies have also shown that women have higher odor intensity ratings compared to men^36^. This may be due to both hormonal and behavioral factors: women tend to express more interest in odors and may be more aware of and attentive to the odors used in the test^17,37^. A gradual decrease in olfactory sensitivity with aging is also well documented^38^. Our findings confirm previous reports of reduced odor intensity and identification in older people compared to young and middle-aged participants^39^ and indicate a degree of unidentified smell loss in our older participants^5^.

We found that heritability was low for odor intensity but moderate for odor pleasantness. These results should be interpreted with caution because our sample size is too small for certainty in the heritability estimates, and dizygotic twins made up only ∼13% of the total twin pairs. Previous studies have found strong effects of heritability on olfactory function, but these effects were specific to individual odors, such as sensitivity to isoamyl acetate^40^ and androstenone^22,40^. The pleasantness of cinnamon is heritable and has been mapped to chromosome 4 through linkage analysis, but this region has no odorant receptor genes^41^. Genetic variation in a single olfactory receptor can affect odor intensity and pleasantness, accounting for person-to-person differences in odor pleasantness^21^. However, many of these strong genetic effects on olfactory function are lost with aging^39^ or when an odor is a mixture of multiple chemicals^42^. Others have found no or only modest genetic effects on detection, intensity, pleasantness, and identification ratings of UPSIT odors^41,43–45^. Overall, as long as a smell test does not include an odor known to have genetic effects on odor pleasantness or detection, and/or includes odors with a mixture of multiple chemicals (as done in UPSIT and SCENTinel)^46^, the metric does not have a large effect on person-to-person differences in test performance.

Several limitations to this study should be noted. First, this study was conducted outdoors, at a festival, resulting in an uncontrolled environment with competing distractions. This likely created more variability that if the data were collected under controlled conditions. Second, all participants included in the analyses were normosmic and accurately completed the SCENTinel^™^ test, but this prevented us from being able to calculate the test-retest reliability and heritability of categorical variables due to low variability in dichotomized (pass/fail) test scores. Lastly, the test-retest reliability was analyzed with data collected on consecutive days; more time between tests would yield more informative results.

In conclusion, a smell test that can serve as a population-wide screen for smell function can help screen for disease, including SARS-CoV-2 infection. SCENTinel^™^ is a rapid, inexpensive smell test that can easily be deployed population-wide. In this study, we found that SCENTinel^™^ reproduces age and sex differences in olfactory function found in other smell tests and had a moderate to high test-retest reliability across two days. We found low to moderate heritability effects on SCENTinel^™^ odor intensity and odor pleasantness subtests, respectively. Future studies will use SCENTinel^™^ with more contrasted distractors to improve odor identification in normosmic individuals. Overall, results from this study support the use of SCENTinel^™^ as a population-wide tool to screen for smell function.

## Data Availability

The data and analysis script will be publicly available on OSF upon acceptance of the publication (https://osf.io/g89dq/).

## Acknowledgements

We would like to thank Dr. Robert Pellegrino, Dr. Ha Nguyen, Dr. May Cheung, Ahmed Barakat, Justin Doran, Dan Doran, Mitchell Krieger, Ke Lin, CJ Arayata, Woody Felice, Lauren Colquitt, Riley Herriman, Sarah Marks, Riley Koch, Ivona Sasimovich, and Aurora Toskala for their assistance in collecting data at the Twins Days Festivals.

## Author Contributions

SRH: Conceptualization, Formal analysis, Investigation, Writing – Original Draft, Writing – Review & Editing, Visualization, Project Administration; CL: Formal analysis, Writing – Original Draft, Writing – Review & Editing, Visualization; MEH: Investigation, Supervision, Project administration; KB: Investigation, Project administration; AH: Investigation, Project administration; PVJ: Writing – Review & Editing, Funding acquisition; VP: Conceptualization, Methodology, Writing – Review & Editing, Funding acquisition; PHD: Conceptualization, Methodology, Writing – Review & Editing, Funding acquisition; and DRR: Conceptualization, Methodology, Investigation, Writing – Review & Editing, Supervision, Project administration, Funding acquisition

