## Supplementary Data for "Low to moderate genetic influences on the rapid smell test SCENTinel™"

### Supplementary Table S1. Participant Demographics in the Test-Retest.

| | N | % |
|---|---|---|
| **Total subjects** | **118** | **100** |
| **Age (median [IQR] years)** | **38 [25-54]** | |
| Young (18-41) | 69 | 58 |
| Middle (42-63) | 36 | 31 |
| Old (64-88) | 13 | 11 |
| **Sex** | | |
| Female | 82 | 69 |
| Male | 36 | 31 |
| Other | 0 | 0 |
| Prefer not to say | 0 | 0 |
| **Race/Ethnicity** | | |
| White | 105 | 89 |
| Black or African American | 3 | 2.5 |
| Asian (including South Asian and Asian Indian) | 2 | 1.7 |
| Hispanic or Latino | 1 | 0.8 |
| American Indian or Alaska Native | 1 | 0.8 |
| Native Hawaiian or Other Pacific Islander | 2 | 1.7 |
| Other | 3 | 2.5 |
| Prefer not to say | 1 | 0.8 |
| **Twin status** | | |
| Monozygous | 82 | 70 |
| Dizygous | 6 | 5 |
| Triplet | 0 | 0 |
| Singleton | 30 | 25 |

**Supplementary Table S2. Components of odors included in the SCENTinel™ 2.1 test.**

| Odor | Components |
|---|---|
| Flower[1] | 2-Phenylethanol [CAS 60-12-8]<br>Benzoic acid<br>Phenyl methyl ester [CAS 120-51-4]<br>Linalool [CAS 78-70-6]<br>Geraniol [CAS 106-24-1]<br>Citronellol [CAS 106-22-9]<br>Nerol [CAS 106-25-2]<br>Geranyl acetate [CAS 105-87-3]<br>Rose oxide L [CAS 16409-43-1]<br>Methyl 2-nonynoate [CAS 111-80-8] |
| Coffee[2] | Vanillin [CAS 121-33-5]<br>Benzaldehyde [CAS 100-52-7]<br>Benzyl benzoate [CAS 120-51-4]<br>gamma-Hexalactone [CAS 695-06-7]<br>Ethyl vanillin [CAS 121-32-4]<br>Ethyl maltol [CAS 4940-11-8]<br>3-Methylbutyraldehyde [CAS 590-86-3]<br>Acetyl propionyl [CAS 600-14-6] |
| Bubblegum[1] | Phenyl methyl ester [CAS 120-51-4]<br>Cyclohexene [CAS 5989-27-5]<br>Vanillin [CAS 121-33-5]<br>Ethyl propionate [CAS 105-37-3]<br>Ethyl butyrate [CAS 105-54-4]<br>beta-Pinene [CAS 127-92-2]<br>Isoamyl acetate [CAS 123-92-2]<br>Myrcene [CAS 123-35-3]<br>Cinnamic aldehyde [CAS 104-55-2] |
| Orange[2] | Limonene [CAS 5989-27-5]<br>Methyl dihydrojasmonate [CAS 24851-98-7]<br>Ethyl methylphenylglycidate [CAS 77-83-8]<br>beta-Ionone [CAS 14901-07-6]<br>4-(4-Hydroxyphenyl)-2-butanone [CAS 5471-51-2]<br>Citral [CAS 5392-40-5]<br>alpha-Methylbenzyl acetate [CAS 93-92-5]<br>Ethyl maltol [CAS 4940-11-8]<br>Allyl heptanoate [CAS 142-19-8]<br>Linalool [CAS 78-70-6]<br>Hydroxycitronellol [CAS 107-74-7]<br>Decanal [CAS 112-31-2]<br>1-Methyl-4-(4-methyl-3-pentenyl)cyclohex-3-ene-1-carbaldehyde [CAS 52475-86-2]<br>Isohexenyl cyclohexenyl carboxaldehyde [CAS 37677-14-8]<br>beta-Myrcene [CAS 123-35-3]<br>Methoxy dicyclopentadiene carboxaldehyde [CAS 86803-90-9]<br>beta-Pinene [CAS 127-91-3]<br>Citronellol [CAS 106-22-9]<br>alpha-Pinene [CAS 80-56-8]<br>1-(2,6,6-Trimethylcyclohexa-1,3-dienyl)-2-buten-1-one [CAS 23696-85-7] |

| | |
|---|---|
| Strawberry[2] | Benzyl benzoate [CAS 120-51-4]<br>Ethyl 2-methyl-1,3-dioxolane-2-acetate [CAS 6413-10-1]<br>Ethyl methylphenylglycidate [CAS 77-83-8]<br>Ethylene brassylate [CAS 105-95-3]<br>gamma-Undecalactone [CAS 104-67-6]<br>Benzyl isobutyrate [CAS 103-28-6]<br>Limonene [CAS 5989-27-5]<br>Benzyl acetate [CAS 140-11-4]<br>Ethyl maltol [CAS 4940-11-8]<br>Ethyl butyrate [CAS 105-54-4]<br>beta-Pinene [CAS 127-91-3]<br>p-Mentha-1,4-diene [CAS 99-85-4]<br>Methyl cinnamate [CAS 103-26-4]<br>Citral [CAS 5392-40-5]<br>alpha-Pinene [CAS 80-56-8]<br>Benzyl salicylate [CAS 118-58-1] |
| Banana[2] | Methyl dihydrojasmonate [CAS 24851-98-7]<br>Isoamyl acetate [CAS 123-92-2]<br>Ethyl vanillin [CAS 121-32-4]<br>Benzyl benzoate [CAS 120-51-4]<br>Vanillin [CAS 121-33-5]<br>p-Methoxybenzaldehyde [CAS 123-11-5]<br>Limonene [CAS 5989-27-5]<br>Piperonal [CAS 120-57-0]<br>Butylated hydroxytoluene [CAS 128-37-0]<br>Ethyl butyrate [CAS 105-54-4]<br>Coumarin [CAS 91-64-5]<br>Benzyl acetate [CAS 140-11-4]<br>Allyl hexanoate [CAS 123-68-2]<br>2-Butanone, 4-(4-hydroxyphenyl)- [CAS 5471-51-2]<br>Ethyl maltol [CAS 4940-11-8]<br>Ethyl methylphenylglycidate [CAS 77-83-8] |
| Woody[2] | Benzyl benzoate [CAS 120-51-4]<br>1-(2,3,8,8-tetramethyl-1,2,3,4,5,6,7,8-octahydronaphthalen-2-yl)ethanone [CAS 54464-57-2]<br>Hexyl cinnamal [CAS 101-86-0]<br>Cedrene [CAS 11028-42-5]<br>Acetyl cedrane [CAS 32388-55-9]<br>Isobornyl acetate [CAS 125-12-2]<br>Cedrol [CAS 77-53-2]<br>Limonene [CAS 5989-27-5]<br>Linalyl acetate [CAS 115-95-7]<br>1-(2-tert-Butyl cyclohexyloxy)-2-butanol [CAS 139504-68-0]<br>Linalool [CAS 78-70-6]<br>Dimethylcyclohex-3-ene-1-carbaldehyde [CAS 68737-61-1]<br>1,2,3,3a,4,5,6,8a-Octahydro-4,8-dimethyl-2-(1-methylethylidene)-6-azulenol [CAS 89-88-3]<br>Methyl atrarate [CAS 4707-47-5]<br>Eucalyptol [CAS 470-82-6]<br>Eugenol [CAS 97-53-0]<br>Benzyl cinnamate [CAS 103-41-3]<br>alpha-Pinene [CAS 80-56-8]<br>Camphene [CAS 79-92-5]<br>beta-Caryophyllene [CAS 87-44-5]<br>Longifolene [CAS 475-20-7] |

| | |
|---|---|
| Coconut[2] | Benzyl benzoate [CAS 120-51-4]<br>gamma-Nonalactone [104-61-0]<br>Methyl dihydrojasmonate [CAS 24851-98-7]<br>Ethyl vanillin [CAS 121-32-4]<br>gamma-Octalactone [CAS 104-50-7]<br>Ethylene brassylate [CAS 105-95-3]<br>1-(2,3,8,8-tetramethyl-1,2,3,4,5,6,7,8-octahydronaphthalen-2-yl)ethanone [CAS 54464-57-2]<br>2-methyl-3-(4-propan-2-ylphenyl)propanal [CAS 103-95-7]<br>2-Isobutyl-4-methyltetrahydro-2H-pyran-4-ol [CAS 63500-71-0]<br>Vanillin [CAS 121-33-5]<br>Coumarin [CAS 91-64-5]<br>Ethyl maltol [CAS 4940-11-8]<br>Linalool [CAS 78-70-6]<br>Piperonal [CAS 120-57-0]<br>gamma-Undecalactone [CAS 104-67-6]<br>Benzyl salicylate [118-58-1] |
| Lemon[2] | Benzyl benzoate [CAS 120-51-4]<br>Citral [CAS 5392-40-5]<br>Methyl dihydrojasmonate [CAS 24851-98-7]<br>Limonene [CAS 5989-27-5]<br>Hexamethylindanopyran [CAS 1222-05-5]<br>Ethyl maltol [CAS 4940-11-8]<br>beta-Pinene [CAS 127-91-3]<br>3,7-Demethyl-2,6-nonadienenitrile [CAS 61792-11-8]<br>p-Mentha-1,4-diene [CAS 99-85-4]<br>Ethylene brassylate [CAS 105-95-3]<br>beta-Myrcene [CAS 123-35-3]<br>alpha-Pinene [CAS 80-56-8]<br>2,6-Demethyl-5-heptenal [CAS 106-72-9]<br>Benzyl salicylate [CAS 118-58-1] |

Odors are from [1]Givaudan, Cincinnati, OH, or [2]Robertet, Mount Olive, NJ.

**Supplementary Table S3. SCENTinel$^{TM}$ overall score algorithms.**

| Scoring response pattern | Detection | Intensity (range: 1-100) | Identification attempt | | Outcome | Probability (chance outcome) |
|---|---|---|---|---|---|---|
| | | | First | Second | | |
| **1** | Correct | ≥ 21 | Correct | NA | ***Pass*** | 0.07 |
| **2** | Correct | ≥ 21 | Incorrect | Correct | ***Pass*** | 0.07 |
| **3** | Correct | ≥ 21 | Incorrect | Incorrect | ***Pass*** | 0.13 |
| **4** | Incorrect | ≥ 21 | Correct | NA | ***Pass*** | 0.13 |
| **5** | Correct | ≤ 20 | Correct | NA | **Fail** | 0.02 |
| **6** | Correct | ≤ 20 | Incorrect | Correct | **Fail** | 0.02 |
| **7** | Correct | ≤ 20 | Incorrect | Incorrect | **Fail** | 0.03 |
| **8** | Incorrect | ≤ 20 | Correct | NA | **Fail** | 0.03 |
| **9** | Incorrect | ≥ 21 | Incorrect | Correct | **Fail** | 0.13 |
| **10** | Incorrect | ≤ 20 | Incorrect | Correct | **Fail** | 0.03 |
| **11** | Incorrect | ≥ 21 | Incorrect | Incorrect | **Fail** | 0.26 |
| **12** | Incorrect | ≤ 20 | Incorrect | Incorrect | **Fail** | 0.07 |

NA, not applicable.
